# Tumor-Specific Decisions Using Tumor-Agnostic Evidence from Basket Trials: A Bayesian Hierarchical Approach

**DOI:** 10.1101/2023.09.19.23295807

**Authors:** Yilin Chen, Josh J. Carlson, Felipe Montano-Campos, Anirban Basu, Lurdes Y.T. Inoue

## Abstract

**Purpose:** Treatment effect heterogeneity across tumor types remains a challenge to evidence interpretation and implementation of tumor-agnostic drugs (TADs), which are typically approved based on basket trial evidence. We sought to use Bayesian hierarchical models (BHM) to assess heterogeneity and improve estimates of tumor-specific treatment outcomes, which are crucial for healthcare decision-making.

**Methods:** We fitted BHMs and Bayesian fixed-effect models to evaluate the objective response rate (ORR), the median progression-free survival (mPFS), and the overall survival (mOS). We estimated the posterior distribution of outcomes for each tumor type, the pooled effects, and intra-class correlations (ICC). Using published basket trial evidence for pembrolizumab (KEYNOTE-158/KEYNOTE-164), we obtained the predictive outcomes in a new cancer type drawn from the same population. In the base case, we assumed non-informative priors with uniform distributions for between-tumor standard deviation. We performed sensitivity analyses with various priors to account for uncertainty in the prior specification.

**Results:** The BHMs shrunk the original tumor-specific estimates toward a pooled treatment effect. The borrowing of information across tumor types resulted in less variability in the posterior tumor-specific estimates compared to the original trial estimates, reflected in narrower 95% credible intervals (CrLs). We found low heterogeneity for ORR but high heterogeneity for mPFS and mOS across cancers (ICC: 0.22, 0.87, 0.7). The predicted posterior means and 95%CrLs were 0.37 (0.15-0.64) for ORR, 3.75 months (0.24-50.45) for mPFS, and 13.76 months (0.42-276.49) for mOS, respectively.

**Conclusions:** Borrowing information through BHM can improve the precision of tumor-specific estimates, thereby facilitating more robust policy decisions regarding TADs. Our analysis revealed high heterogeneity and uncertainty in survival endpoints. Both pooled and tumor-specific estimates are informative for clinical and coverage decision making.

**Highlights:**

- Bayesian hierarchical models could enhance precision and reduce uncertainty of estimates derived from basket trial evidence, potentially improving confidence in tumor-agnostic decision making, despite small sample sizes in some tumor types.
- Our study highlights high variability in treatment effects of pembrolizumab across tumor types with respect to survival endpoints, although treatment effects appear more consistent when judged by objective response rate at approval. Understanding heterogeneity in treatment effects following accelerated approvals based on surrogate endpoint is crucial for clinical and coverage decision making.
- This article demonstrates the use of Bayesian methods to estimate posterior distributions of tumor-specific and aggregated treatment effects (ORR, median PFS, and median OS) from basket trials. Choosing between fixed-effect or random-effects model to evaluate pooled treatment effects depends on the level of heterogeneity in effect sizes across tumor types.

## Introduction

Precision medicine has enabled the development of drugs that target specific molecular targets. In oncology, tumor-agnostic drugs (TADs) have begun to shift the therapeutic paradigm from organ-based treatment to a molecular target-based approach.^1–3^ In 2017, the Food and Drug Administration (FDA) granted an accelerated approval for the first TAD, pembrolizumab, for the treatment of adult and pediatric patients with unresectable or metastatic, microsatellite instability–high (MSI-H) or mismatch repair–deficient (dMMR) solid tumors that have progressed after prior treatment.^4^ As of 2022, FDA has approved five medicines for tumor-agnostic indications, with many more in the drug development pipeline.^5–8^ This paradigm shift in drug development has driven the field of precision medicine to consider the molecular details of cancer biology to identify opportunities to improve the treatment of patients with cancer.^9, 10^ Histology-independent treatments have the potential to provide substantial benefits to patients who currently have few or no therapeutic alternatives.

The accelerated approvals for pembrolizumab and other TADs were based on basket trials characterized by a small number of multi-cohort, single-arm studies.^11^ Basket trials simultaneously evaluate the effect of a targeted agent in various disease subtypes that harbor the same genetic or molecular alteration.^12^ Planning a basket trial is administratively more efficient than conducting separate trials for each cancer type. It has predominantly been used for early-phase oncology drug development, with a recent review finding 37 ongoing basket trials registered in ClinicalTrials.gov.^2, 13^ The emergence of TADs and basket trials has created substantial innovation and unique challenges for drug approval and value assessment for regulatory and health technology assessment (HTA) agencies. One major concern of analyzing early phase basket trials is the potential for heterogeneity of the treatment effect in various tumor types.^14, 15^ For example, the Phase II KEYNOTE-158 study enrolled 233 patients with advanced MSI-H/dMMR cancers and 37 different tumor types previously treated.^16, 17^ The study showed the objective response rate (ORR) ranging from 18.2% in pancreatic cancer to 48% in endometrial cancer. Moreover, even though it is approved for use in all solid tumors, it is difficult to extrapolate these results to a tumor type not included in the basket trial with the same biomarker or genetic mutation.

Bayesian hierarchical models (BHM), or Bayesian random-effects models, are particularly well-suited for handling issues related to heterogeneity and information from varying sample sizes. They are adept in borrowing information on treatment effects across tumors, thereby increasing the precision of tumor-specific estimates while down-weighting extreme results in cohorts with small sample sizes.^18–20^ It has advantages over conventional stand-alone subgroup analyses (also known as the approach of no borrowing), which often lack sufficient power to detect treatment effects because of the small sample size of each stratum.^21^ At the other extreme, a simple pooled analysis, i.e., complete pooling, would not account for systematic patient-to-patient differences across tumor types.^19, 22–24^ BHMs are unique in that they are able to simultaneously account for heterogeneity in treatment outcomes between tumor types and patient-to-patient variability. The uncertainty in between-tumor heterogeneity can also be directly modeled.^25^ BHMs are justified under the assumption of exchangeable treatment effects, which means that it is unknown *a priori* which subgroups perform better than others or that the subgroups are drawn from the same underlying overall population.^26^

BHMs have served an important role in multiple settings in health sciences research, including basket trial design, trial efficacy evaluation, observational studies, comparative effectiveness research, meta-analysis, and evaluation of disease subtypes.^27–31^ Studies using BHMs have only evaluated ORR and no other commonly used primary endpoints in clinical trials, such as median progression-free survival (mPFS) and median overall survival (mOS). The objective of this study was to estimate the treatment outcomes for each tumor type based on data from the KEYNOTE-164 [NCT02460198] and KEYNOTE-158 [NCT02628067] trials, using Bayesian random-effects models.^16, 17, 32^ Compared to a random-effects model that allow the true effect sizes to differ, a fixed-effect model assumes that there is one true effect size, and that all difference in observed effects are due to within-group error, and thus no between-group variance.^33^ We implemented BHMs and Bayesian fixed-effect models to obtain the tumor-specific and pooled treatment effects across all tumor types, and the predictive distributions in an unrepresented, new cancer type from the same overall population. The study findings will provide improved tumor-specific estimates for pembrolizumab using early-phase basket trial evidence and insights into the methods to consider when assessing the clinical benefits of TADs. The methods presented in this study can be applied to the future assessment of TADs.

## Methods

### Overview of study methods and outcomes

We fitted three types of models to evaluate the ORR, mPFS, and mOS: (1) Bayesian hierarchical model, which assumes varying true effect sizes, (2) Bayesian fixed-effect model, which assumes one true effect size, and (3) Bayesian fixed-effects model, assuming no between-tumor variation but with varying mean effects (hence the plural term fixed-effects). Posterior distributions summarized with the posterior mean, median, and 95% posterior credible intervals (CrLs) were obtained for treatment effects in each tumor type. The posterior distribution can be expressed as: *p*(*θ*_1_, …, *θ*_*N*_, *ϕ*|*Data*) =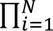 *p*(*Data*_*i*_|*θ*_*i*_, *ϕ*)*p*(*θ*_*i*_|*ϕ*)*p*(*ϕ*). *Data_i_* refers to data from tumor site i and *Data* refers to the data from all tumor sites.

Metrics of heterogeneity were calculated to quantify total variation across tumor types.^25^ Posterior distributions for the between-tumor standard deviation (SD) and 95% CrLs were computed and summarized. Intra-class correlation (ICC) provides a measure of the proportion of total variation that is explained by between-tumor variability, expressed as 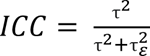. ICC is the ratio of the between-cluster variance (τ^2^) to the total variance (τ^2^ + 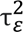), which ranges from 0 to 1.^34, 35^ Therefore, low ICC indicates less variation in treatment effects between tumor types, which means a tumor-aggregated decision may be more appropriate, and vice versa.

### Setting

Pembrolizumab was approved for treating previously treated adult and pediatric patients with unresectable or metastatic MSI-H/dMMR (i.e., MSI-H in this paper) cancers based on tumor response rate and durable clinical benefit in two Phase II studies. A total of 63 patients (cohort B) with MSI-H colorectal cancer (CRC) and 233 patients (cohort K) with MSI-H non-CRC were included in this analysis. The data cut-off date was September 4, 2018 for KEYNOTE-164 and October 5, 2020 for KEYNOTE-158 studies.^16, 17, 32, 36^ The ORR, mPFS, and mOS data for eight tumors (colorectal, endometrial, gastric, cholangiocarcinoma, pancreatic, small intestine, ovarian, and brain) are summarized in Table 1. Due to the nature of the study, which involved no direct contact with human participants, the ethical approval was not obtained.

**Table 1.** Trial-reported clinical outcomes by tumor types.

| **Tumor type** | **Number of patients** | **ORR, % (95%CI)** | **Median PFS, months, (95%CI)** | **Median OS, months, (95%CI)** |
| --- | --- | --- | --- | --- |
| Colorectal | 63 | 33 (22, 46) | 4.1 (2.1, 18.9) | 33* (19.2, NR) |
| Endometrial | 79 | 48.5 (36.2, 61) | 13.1 (4.3, 34.4) | 46.5* (27.2, NR) |
| Gastric | 42 | 31 (17.6, 47.1) | 3.2 (2.1, 12.9) | 11 (5.8, 31.5) |
| Cholangiocarcinoma | 22 | 40.9 (20.7, 63.6) | 4.2 (2.1, 24.9) | 19.4 (6.5, NR) |
| Pancreatic | 22 | 18.2 (5.2, 40.3) | 2.1 (1.9, 3.4) | 3.7 (2.1, 9.8) |
| Small intestine | 25 | 48 (27.8, 68.7) | 23.4 (4.3, NR) | NR (16.2, NR) |
| Ovarian | 24 | 33.3 (15.6, 55.3) | 2.2 (2.0, 6.2) | 33.6 (11.0, NR) |
| Brain | 13 | 0 (0, 24.7) | 1.1 (0.7, 2.1) | 5.6 (1.5, 16.2) |

Abbreviations: CI, confidence interval; ORR, objective response rate; PFS, progression-free survival; OS, overall survival; NR, not reached at the end of study follow-up period.
\* Estimated from the fitted exponentiation distribution to the digitized Kaplan-Meier curve data.

### Model specification

#### Objective response rate

We fitted a Bayesian hierarchical model to the ORR data. The first unit of the hierarchical model is the patient, and the second unit of analysis is the tumor type, with patients grouped or nested within tumor types. For each tumor type, the observed response rates are modeled using a binomial distribution, where *y*_*j*_ is the number of responses for tumor j, *n*_*j*_ is the number of patients for tumor type j, and *p*_*j*_ is the probability of response. A logit transformation is applied to the probability of response, expressed as *θ*_*j*_ =l n (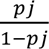), and is assumed to be normal. The hyperparameters are the overall mean log-odds of response (*ϕ*), average across all possible tumor types, and the between-tumor SD (*τ*). The two hyperparameters are treated as unknown random variables. Specifically, we assumed the following probability distributions.

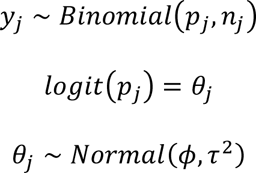

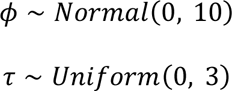

In sensitivity analysis, we repeated our analysis using other prior distributions for the square root of the hypervariance (*τ*): (i) the exponential (0.3) distribution (that is, exponential with rate 0.3), another common non-informative prior distribution but with longer tail compared to a uniform distribution; (ii) a truncated normal (0, 100)T(0,) distribution (100 is the variance), which restricted all values to be positive; (iii) a uniform (0, 5) distribution, which assumes equally likely values between 0 and 5; and (iv) an exponential (1) distribution.

All tumor types were included in the analysis except for brain tumor due to the extremely small sample size (i.e., 13 patients).

#### Median Progression-free Survival and Median Overall Survival

Two hierarchical models were fitted separately to model mPFS and mOS. Data from all eight tumor types were included. Due to the positive nature of survival endpoints, we performed a log transformation to median PFS and OS data in the base case and assumed normal distribution for the log-transformed data. The model structure of mPFS and mOS are outlined below, where *y*_*j*_ is the log-transformed mPFS or mOS for tumor j and is assumed to be normal.

Due to the lack of individual-level survival data, we computed the within-tumor variance using the following methods. Note first that under an exponential distribution for the PFS and OS at the individual level data in each tumor type with parameter *λ* (with density function *f*(*x*; *λ*) = *λ* exp(−*λx*)), we can re-express this parameter considering the median survival as 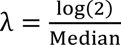. According to Laplace, the distribution of the sample median from a population with a density function *f*(*x*) is asymptotically normal with variance 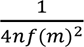, where m is the median and n is the sample size.^37, 38^ Thus, we were able to compute the sample variance for each tumor type by using this result. Finally, we use the delta method to obtain the variance of log (*y*_*j*_), which were denoted as 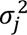 in the model specification above.

The hyperparameters are the overall mean log-mPFS or log-mOS (*ϕ*), average across all possible tumor types, and the between-tumor SD (*τ*). The two hyperparameters are assumed to be normally and uniformly distributed, respectively.

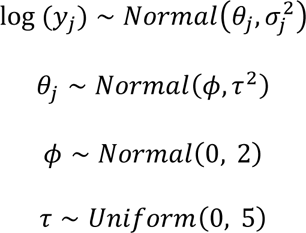

Due to unobserved median OS among several tumor types by the end of the study period, our model also accounted for right-censored observations. This required us to assume that the unobserved outcome value would be greater than a certain value (e.g., end of the study period). In addition, we initialized our Markov-chain Monte Carlo (MCMC) algorithms by setting a value that was larger than the censoring limit.

To define the prior distributions of parameters *ϕ* and τ, we assume a normal distribution for *ϕ* and uniform distribution for τ in base-case analyses. In addition, sensitivity analyses assumed the following distributions for τ: exponential, uniform, and half-normal distributions.^34, 39^ A smaller value of τ implies that the parameters are more similar and results in greater shrinkage, and vice versa.^40^ The priors were selected to be relatively non-informative in the base case to allow the data to primarily determine the degree of similarity among the different tumor types. Weakly informative priors based on scientific knowledge were explored in the sensitivity analyses.

#### Intra-class Correlation

Different methods were used to compute ICC for ORR and survival endpoints. To compute ICC for ORR, we adopted the analysis of variance (ANOVA) estimator approach, which appropriately accounted for the variable size of clusters.^41, 42^ The ANOVA estimator is given by 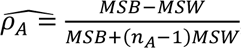, where MSB is the deviations from the group mean and the grand mean and MSW is the deviations of the individual scores from the group mean. Suppose that there are k clusters and that the i^th^ cluster has n_i_ individuals. Then

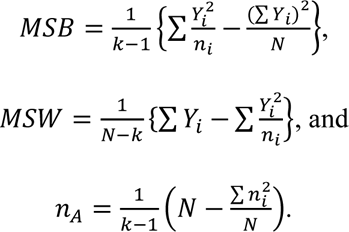

where *Y*_*i*_ = ∑ *X*_*i*_ is the total number of successes in cluster i, and *N* = ∑ *n*_*i*_ is the total number of observations in the study.^41, 42^ For mPFS and mOS models, between-tumor variance was directly obtained from the posterior summaries after applying the exponentiation to transform back the posterior distributions to their original scales. Within-tumor variance was estimated using the observed data and the number of patients as described in the above sections.

### Bayesian Estimation and Inference

A MCMC algorithm was used to generate samples from the posterior distribution, using the likelihood and priors as discussed in previous sections. For each model, 3 parallel chains containing 65,000 samples from the posterior distribution were obtained after a burn-in of 5,000, using a thinning factor of 5. Convergence was assessed using the Gelman-Rubin statistic which evaluates MCMC convergence by analyzing the difference between multiple Markov chains. We also inspected the trace plots and autocorrelation plots for diagnostic checks. The autocorrelation measures how linearly correlated the current value of the chain is to the past values. In addition, prior predictive checks were conducted to assess the appropriateness of the priors and the uncertainty. All Bayesian analyses were performed using R (version 4.1.2) and the package R2JAGS.

## Results

A total of 290 patients with eight types of metastatic or advanced cancers were included in the analysis, including endometrial (n=79), colorectal (n=63), gastric (n=42), small intestine (n=25), ovarian (n=24), cholangiocarcinoma (n=22), pancreatic (n=22), and brain cancers (n=13). The clinical outcomes of ORR, mPFS, and mOS from the KEYNOTE-164 and KEYNOTE-158 trials were summarized in **Table 1**. We reported the mean and 95%CrLs for the posterior distribution of parameters of interest across three outcomes from **Figure 1** to **Figure 4**. The posterior distributions for tumor-specific and pooled outcomes were plotted in **Figure 5**.

**Figure 1.**
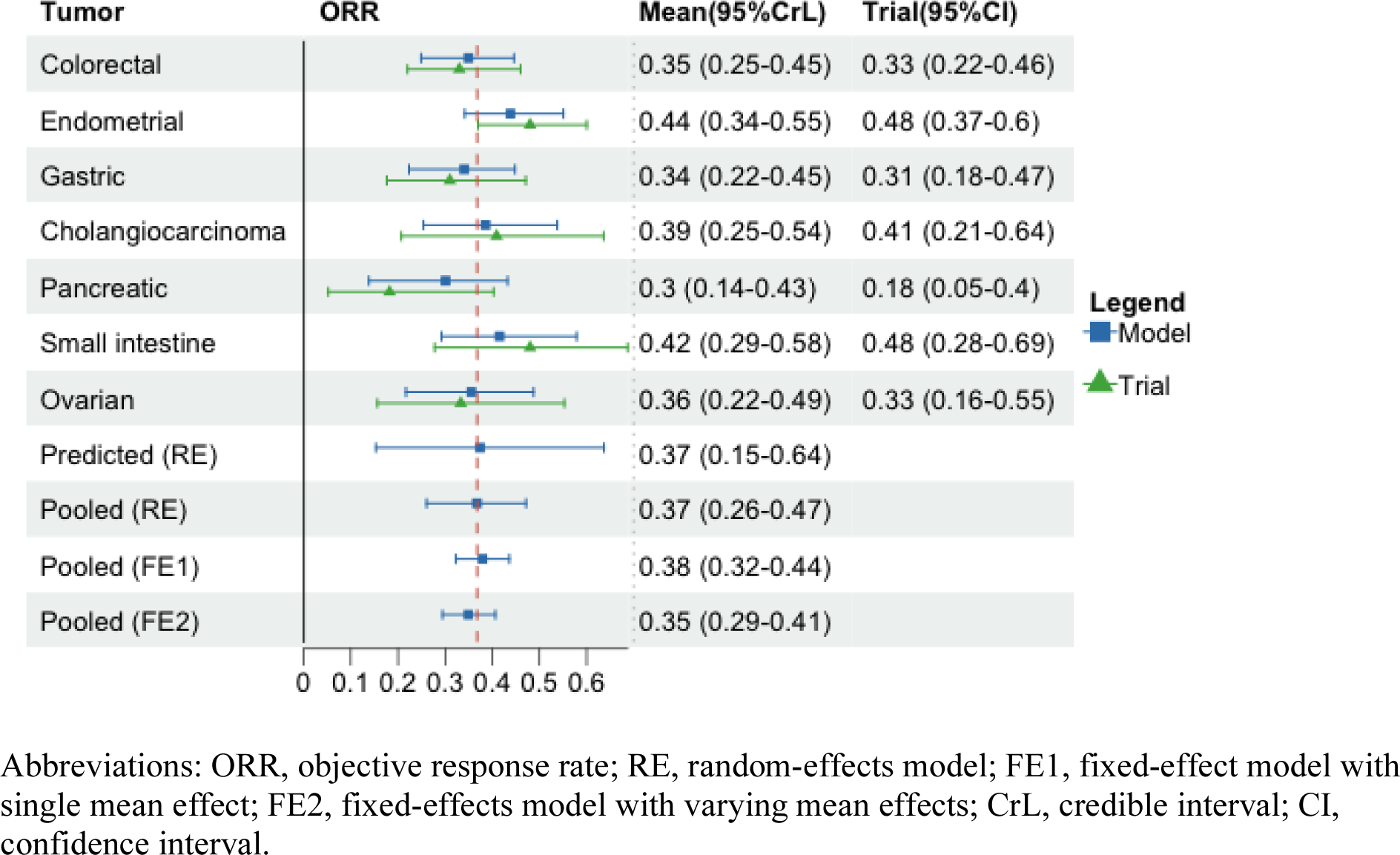
Summary of posterior distributions by tumor types for objective response rate.

**Figure 2.**
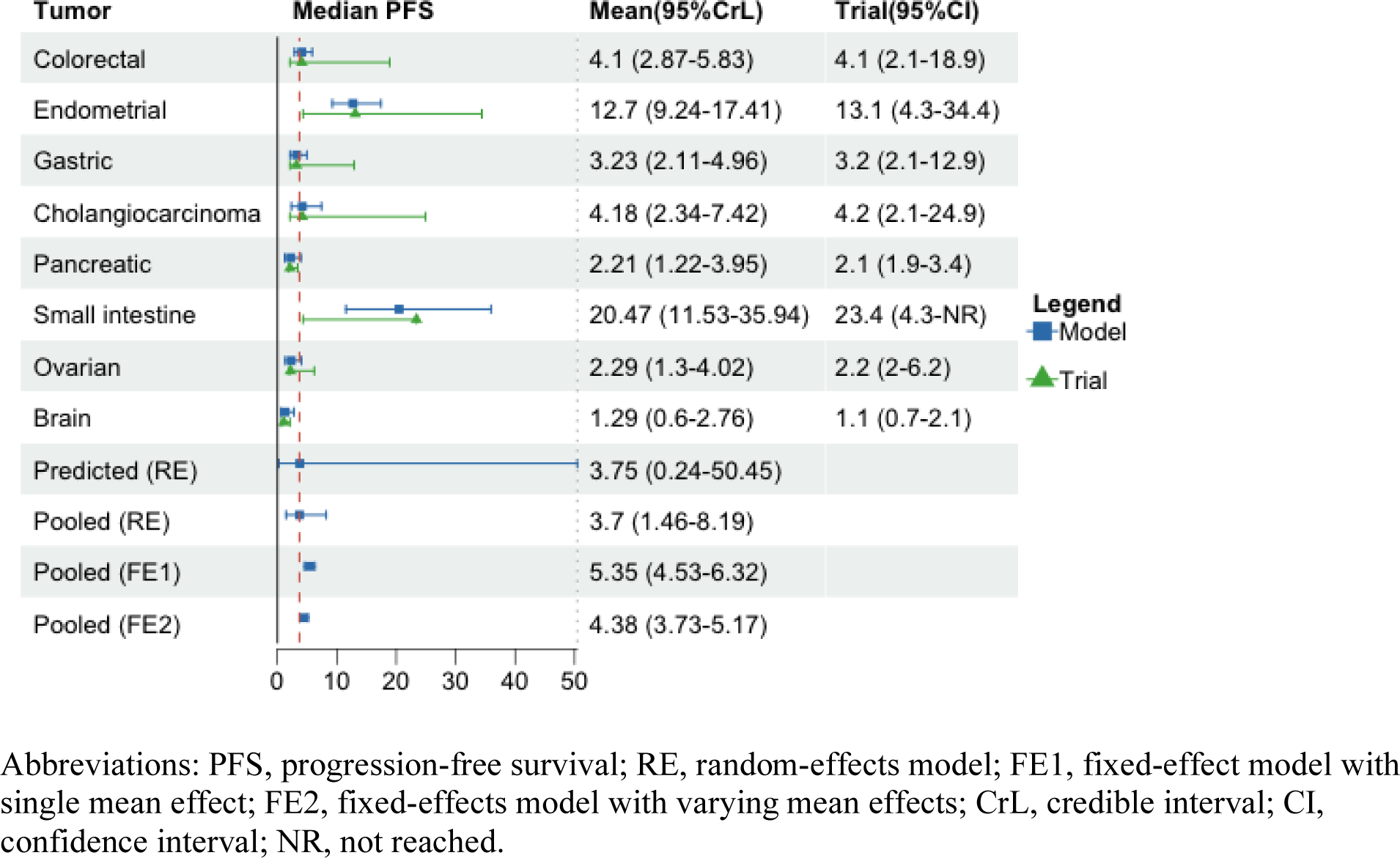
Summary of posterior distributions by tumor types for median progression-free survival.

**Figure 3.**
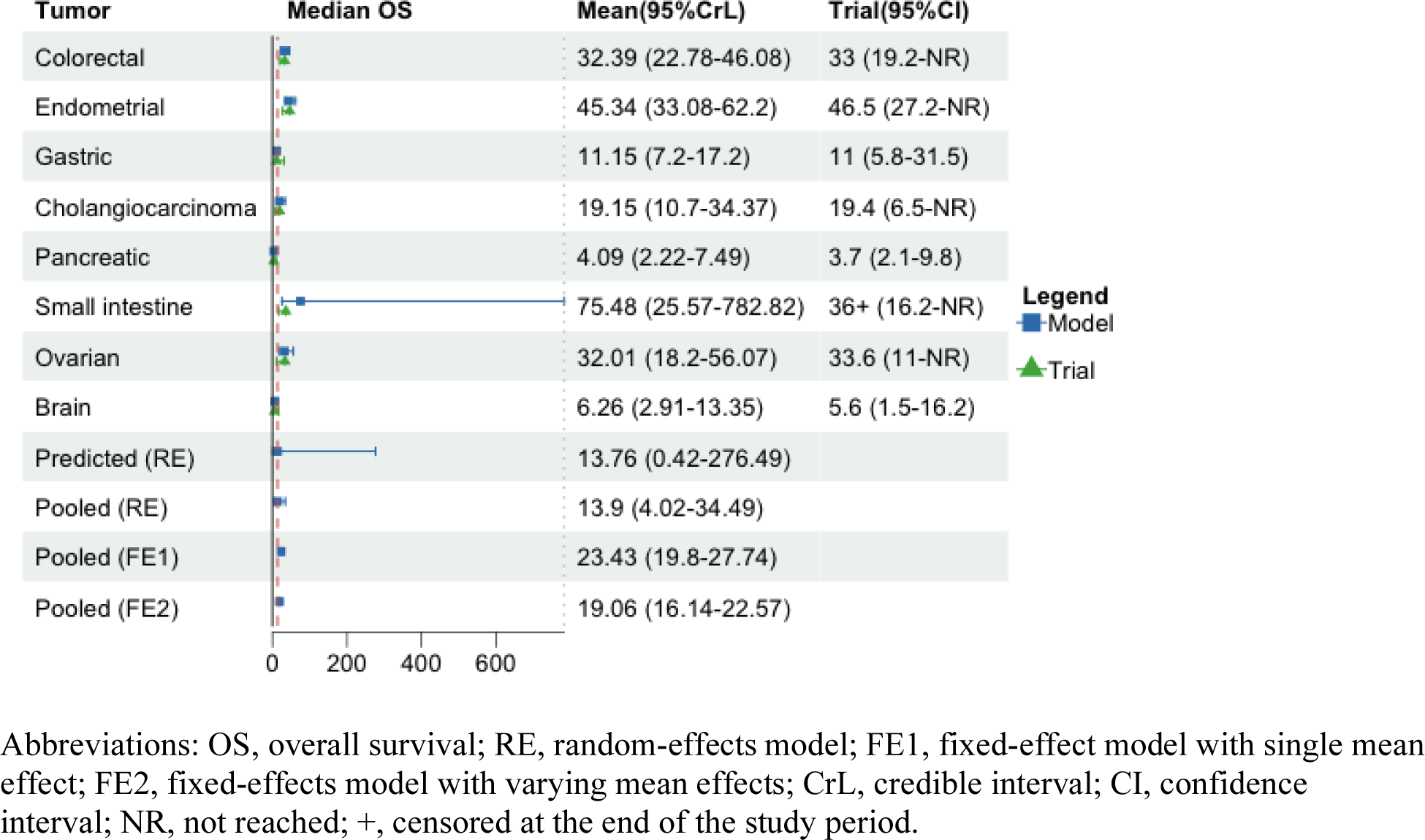
Summary of posterior distributions by tumor types for median overall survival.

**Figure 4.**
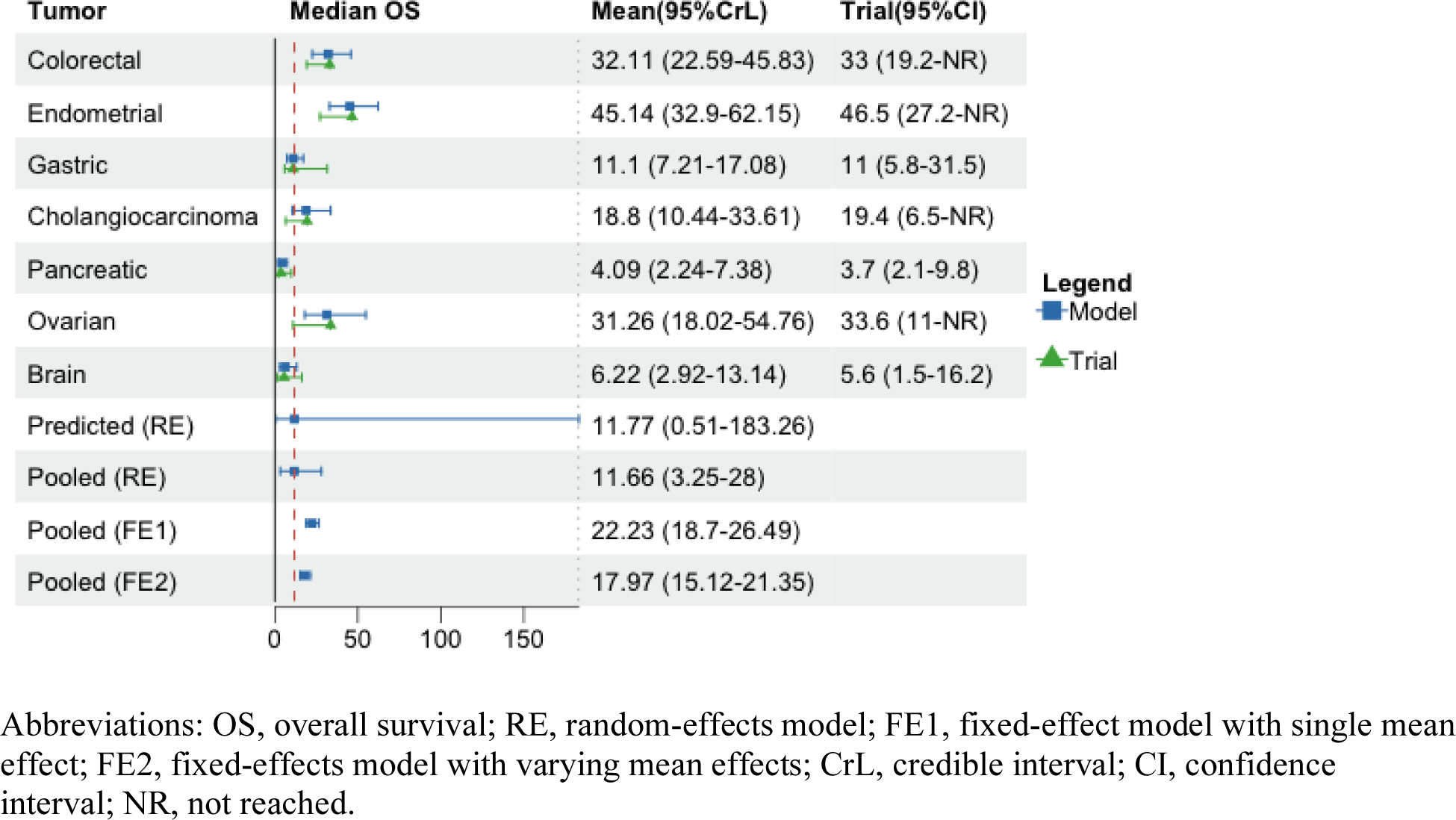
Summary of posterior distributions by tumor types for median overall survival, excluding small intestine cancer.

**Figure 5.**
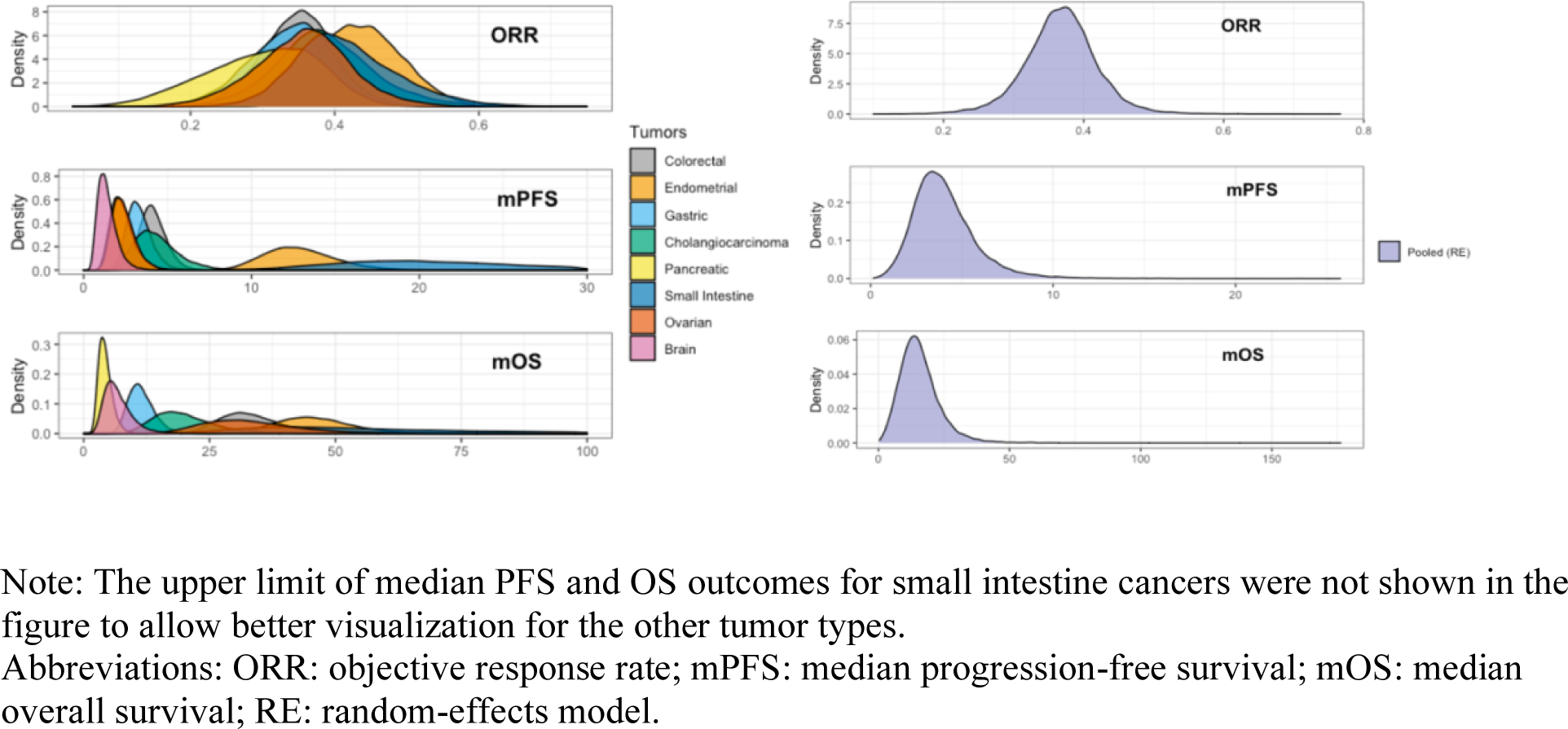
Posterior density plots by tumor types and treatment outcomes.

### Posterior Distributions of Objective Response Rate

The highest mean posterior ORR was observed in endometrial (0.44, 95%CrL: 0.34-0.55), followed by small intestine (0.42, 95%CrL: 0.29-0.58), cholangiocarcinoma (0.39, 95%CrL: 0.25-0.54), ovarian (0.36, 95%CrL: 0.22-0.49), colorectal (0.35, 95%CrL: 0.25-0.45), gastric (0.34, 95%CrL: 0.22-0.45), and pancreatic cancers (0.30, 95%CrL: 0.14-0.43). Using the random-effects model, the pooled mean posterior ORR across seven cancers was 0.37 (95%CrL: 0.26-0.47) and the predictive probability of response in a new, unrepresented MSI-H cancer was 0.37 (95%CrL: 0.16-0.63). Similar mean posterior ORR was obtained from the fixed-effect models with narrower 95% credible intervals (fixed-effect model with single mean effect: 0.38, 95%CrL: 0.32-0.44; fixed-effects model with varying mean effects: 0.35, 95%CrL: 0.29-0.41) (**Figure 1**).

The sensitivity of the BHM results was assessed using alternative prior distributions. The models rendered similar estimates to those obtained with a uniform (0, 3) distribution in the base case. The results showed low estimated heterogeneity between tumors irrespective of the prior distribution, with the intraclass correlation being 0.02 in **eTable 1**.

### Posterior Distributions of Median Progression-free Survival

When different types of tumors were compared for the mean mPFS, small intestine cancer had the longest posterior mean mPFS at 20.47 months (95%CrL: 11.53-35.94), followed by endometrial cancer (12.7, 95%CrL: 9.24-17.41), cholangiocarcinoma (4.18, 95%CrL: 2.34-7.42), colorectal (4.1, 95%CrL: 2.87-5.83), gastric (3.23, 95%CrL: 2.11-4.96), ovarian (2.29, 95%CrL: 1.3-4.02), pancreatic (2.21, 95%CrL: 1.22-3.95), and brain cancers (1.29, 95%CrL: 0.6-2.76). The pooled mean mPFS was 3.7 months (95%CrL: 1.46-8.19) with the random-effects model, with the predictive mean of 3.75 months (95%CrL: 0.24-50.45). The fixed-effect model assuming one mean effect across tumor types yielded higher pooled estimates than those obtained from the fixed-effect model assuming varying mean effects (5.35, 95%CrL: 4.53-6.31; vs. 4.38, 95%CrL: 3.73-5.17) (**Figure 2**).

The sensitivity of the BHM results was assessed by assuming various prior distributions for between-tumor variance (*τ*). The models rendered similar tumor-specific and pooled estimates to those obtained when assuming a uniform distribution (0, 5) in the base case. The results showed high estimated heterogeneity between tumors irrespective of the prior distribution. This can be seen in the intraclass correlation value (0.87) in **eTable 1** and the considerably wide 95%CrLs around the predicted estimate (**Figure 2**).

### Posterior Distributions of Median Overall Survival

Small intestine cancer was estimated to have the longest posterior mean mOS at 75.48 months with wide 95% credible intervals (25.57-782.82) due to its right censoring at the end of study period. The posterior mean mOS was 45.34 months for endometrial cancer (95%CrL: 33.08-62.2), 32.39 months for colorectal cancer (95%CrL: 22.78-46.08), 32.01 for ovarian cancer (95%CrL: 18.2-56.07), 19.15 for cholangiocarcinoma (95%CrL: 10.7-34.37), 11.15 for gastric cancer (95%CrL: 7.2-17.2), 6.26 for brain cancer (95%CrL: 2.91-13.35), and 4.09 for pancreatic cancer (95%CrL: 2.22-7.49). The pooled mean mOS was 13.9 months (95%CrL: 4.02-34.49) with the random-effects model, with the predictive mean of 13.76 months (95%CrL: 0.42-276.49). Similar to mPFS models, the fixed-effect model with a single mean effect across tumor types provided higher pooled estimates than those obtained from the fixed-effect model with varying mean effects (23.43, 95%CrL: 19.8-27.74; vs. 19.06, 95%CrL: 16.14-22.57) (**Figure 3**). The posterior distributions obtained from models excluding small intestine cancer were summarized in **Figure 4**, with similar tumor-specific estimates but lower pooled mean effects compared to models with small intestine cancer.

Sensitivity analysis results were summarized in **eTable 4**. When compared to the base case model, some differences were manifested in the posterior mean estimates for small intestine cancer, the predicted and pooled outcomes, indicating that using BHM for evaluating median OS with censored observations is sensitive to the distribution of between-tumor variance under this setting with limited information about the censored data. The results also showed high estimated heterogeneity between tumors irrespective of the distribution of the data and the prior. The intraclass correlation value was 0.7 (**eTable 1**) and the 95%CrLs around the predicted estimate were wide in both models with and without small intestine cancer (**Figure 3; Figure 4**).

### Model diagnostics

Gelman-Rubin diagnostics estimates were near 1 for all parameters in all models. The trace plots or autocorrelation plots did not suggest convergence failure of the MCMC algorithm.

## Discussion

In this study, we demonstrated the use of Bayesian methods to improve tumor-specific estimates of pembrolizumab using phase II basket trial evidence and obtain posterior distributions of pooled and predicted outcomes, including ORR, median PFS, and median OS. The revised estimates demonstrated markedly less variability than the original estimates for all outcomes, with narrower 95% credible intervals compared to the original CIs (the original trial estimates were obtained using frequentist approach). We also found high heterogeneity in median PFS and median OS across eight tumor types and low heterogeneity in ORR across seven tumor types, excluding brain tumor. The BHM approach shrunk the original tumor-specific ORR estimates towards a pooled treatment effect and the resultant estimates in all tumor types were above 30%. This value is greater than the commonly used regulatory target of 30% with tumor shrinkage used for single-agent anticancer therapies to demonstrate breakthrough activity and for monitoring in phase II basket trials.^43, 44^ The pooled ORR across seven tumor types was 35%-37% using Bayesian random-effects and fixed-effect models, meaning that at least 35% of cancer patients had tumor shrinkage. The BHM approach allows for prediction of outcomes in tumor types outside of the studied types; however, the wide 95%CrLs around these predicted estimates indicate substantial uncertainty, especially for the predicted median PFS and median OS estimates.

### Implications

Our study demonstrated that a Bayesian hierarchical approach could reduce the uncertainty in estimates from available evidence, providing more confidence in tumor-agnostic decision making, given the small sample sizes in some tumor types. This flexible approach can incorporate censored observations of survival endpoints available at trial completion. Moreover, metrics such as ICC can be used to quantify the variation between groups, which can inform a recommendation of pembrolizumab for use in all tumor types or a restricting subset of patients. In addition, BHM can be used to inform many different types of research questions. It has been used in meta-analysis and economic evaluation of healthcare interventions.^40, 45^ For instance, Kwok et al. demonstrated the use of BHM to incorporate all available information from multiple sources, including a meta-analysis of immunosuppressive therapy in idiopathic dilated cardiomyopathy using data from related trials, and a subgroup analysis of the National Institute of Neurological Disorders and Stroke intravenous tissue plasminogen activator stroke trial. Future TAD assessments can benefit from applying Bayesian hierarchical models, such as estimating the posterior probability that patients with each type of cancer demonstrate the largest treatment effect among all cancer patients or the probability of response rate higher than a certain threshold. Such information would provide additional evidence on tumor-specific treatment outcomes, which would inform the approval and coverage decisions.

A high ORR with a long duration of response has been used for accelerated approvals under the assumption that they are highly predictive of longer-term survival outcomes. Our analyses did not support this assumption for pembrolizumab. Specifically, even though the posterior mean ORR in all tumor types was above 30%, we observed low mPFS and mOS in certain tumor types, such as pancreatic and brain tumors. Furthermore, although homogenous ORR outcomes were shown across tumor types, there was high variability for the survival endpoints (i.e., median PFS and OS). In this context, confirmatory trials or post-marketing evidence generation should be considered following accelerated approvals, and could be targeted towards the tumor types with the greatest discordance and/or uncertainty. Future studies can model the correlation of median PFS and OS outcomes in the BHM and explore additional survival endpoints such as 12-month/24-month survival rates to generate more evidence regarding long-term treatment efficacy. In addition, alternative approaches that allow tailored or partial borrowing across strata can be employed when there is high between-tumor heterogeneity in the basket trial setting.^46^

### Considerations for choosing random-effects vs fixed-effect models

Similar to a meta-analysis, we derived the distribution of the pooled treatment outcomes to quantify the overall clinical benefit across tumor types. We explored both Bayesian random-effects and fixed-effect models to obtain a pooled effect size across subgroups, and thus, it is crucial to determine the optimal choice of model for a given scenario. The typical fixed-effect model assumes that there is no between-tumor heterogeneity, or that patients with different tumors are part of a homogenous population, and the only cause for differences in observed effects is the sampling error of tumor sites. To relax the assumption of one true effect size, we further employed a fixed-effects (plural term) model which allows some variation in effect sizes (i.e., FE2 in Figure 1-4). In contrast, the random-effects model has the advantage of assuming there is a pre-specified distribution of true effect sizes and can explicitly quantify between-tumor heterogeneity (τ). The caveat is that this assumption may fail if an outlying group is inconsistent with a certain type of distribution for the between-group variability, potentially resulting in biased estimates for that outlying group. Therefore, it is important to consider sensitivity analyses that verify the robustness of the results to changes in the choices of prior distributions.

The selection of a fixed-effect or a random-effects model depends on how heterogeneous the effect sizes are across tumor types. For example, the fixed-effect pooled estimate of the ORR model might be closer to the true effect size if we believe that patients with different tumor types but sharing the same molecular biomarker respond similarly to the pembrolizumab. However, the assumptions of the fixed-effect model may seem too simplistic in many real-world populations and for long-term outcomes. When there is high heterogeneity across subgroups, such as median PFS and OS models, it may be more plausible to use the pooled effect size from a random-effects model rather than a fixed-effect model.

### Degree of shrinkage

The original tumor-specific estimates were shrunken towards a pooled treatment effect in BHMs which is consistent with prior studies. For example, Chugh and others evaluated the ORR of a phase II trial of Imatinib in 10 histologic subtypes of sarcoma using a BHM. The posterior estimates based on the BHM shrunk the estimates of the subgroup-specific ORR toward the population mean, thus reducing extreme estimates for subtypes with few patients.^47^ We found that greater shrinkage of posterior estimates toward the overall mean occurred for tumor subgroups with relatively smaller sample sizes or less precision. For example, posterior mean ORR of ovarian cancer (raw: 0.33 vs revised: 0.36; n=24 patients) has shrunken to a greater degree than that of colorectal cancer (raw: 0.33 vs revised: 0.35; n=63 patients). In other words, because there is fewer underlying data (i.e., when there is greater uncertainty), the ovarian cancer estimate borrows more information from the other tumor subgroups than the colorectal cancer estimate. In addition, the degree of shrinkage also depends on how far the estimate is from the overall mean. For example, more shrinkage was observed for cholangiocarcinoma (raw: 0.41 vs. revised: 0.39) than for pancreatic cancer (raw: 0.18 vs. revised: 0.3), with equal sample size (n=22 patients). Similar findings are shown in mPFS and mOS models. Prior studies have demonstrated that shrinkage and borrowing of information across subgroups can improve the accuracy of estimates overall than if we were to use the naïve estimates from individual subgroups.^21, 48^

### Prior distributions

Between-tumor variance (*τ*^2^), the shrinkage parameter, estimates the variation across subgroups and controls the strength of information borrowing, was treated as an unknown parameter following a non-informative prior in our study. A small value of *τ*^2^ indicates less heterogeneity across subgroups and thus induces strong information borrowing, whereas a large value of *τ*^2^ induces little information borrowing.^49^ Therefore, it is important to consider prior distributions for *τ*^2^. In our study, we chose non-informative uniformly distributed priors in the base case to allow data rather than prior knowledge to primarily determine the degree of borrowing. This is consistent with previous investigation around the priors in the literature. Furthermore, when more prior information is desired, for instance to restrict *τ* away from very large values, Gelman et al. recommended working within the half-t family of prior distributions, which are more flexible. However, the inverse-gamma family of non-informative prior distributions is not recommended due to the sensitive resulting inferences when *τ* is estimated to be near zero.^34, 39^

In a fully Bayesian approach, the third level of the hierarchical model consists of the prior distributions of the hyperparameters which quantify knowledge based on external information. Vague, non-informative prior distributions may be used in the absence of relevant external information, or to permit inferences to depend exclusively on the present data.^40^ In an empirical Bayes approach, in contrast, the hyperparameters are derived from the data and then treated as known values.^50^ However, this often results in overly precise estimates of the first-level parameters as it ignores the uncertainty in the values of the hyperparameters.^51^

### Limitation

Our study is not without limitations. The main assumption of this approach is exchangeability that efficacy is similar across baskets, with different tumors not determining a particular ordering of effectiveness a priori. It is unknown whether the exchangeability assumption will apply to unrepresented tumors. Furthermore, our results may be dependent on the data distribution and the priors. To address the uncertainty, we used the existing literature and knowledge to come up with prior distributions that make clinical sense and tested a few to examine whether the results change significantly. In the absence of individual patient data, we computed the within-tumor variance for mPFS and mOS models using above-mentioned methods. We also used published Kaplan-Meier curves to estimate median OS for colorectal and endometrial cancers, assuming an exponential distribution. The assumption of exponential distribution for time-to-event outcomes is also a potential limitation since it is assuming constant hazard. Our proposed model, however, could be extended to consider other parametric distributions such as the Weibull distribution and eliciting other quantiles of the Kaplan-Meier curves instead of just the median OS.

## Conclusion

Assuming exchangeable effects of tumor-agnostic therapies across cancers, BHM can be useful for studying treatment effects by reducing the variability around estimates and improving precision in estimates overall through borrowing of information, compared to stand-alone subgroup analyses with limited sample sizes in some strata. Our findings suggest that making tumor-agnostic drug indication and coverage decisions simply based on the pooled treatment outcomes in a basket trial may not be appropriate in some tumor types. Both pooled and tumor-specific ORR, mPFS, and mOS posterior estimates may be useful in informing decision making. Finally, this approach permits quantifying the between-tumor heterogeneity across subgroups, characterizing the proportion of variance explained by the between-tumor variability, and offers further evidence on survival endpoints in an early-phase basket trial.

## Data Availability

All data produced in the present work are contained in the manuscript

