## Supplemental Materials for "Tumor-Specific Decisions Using Tumor-Agnostic Evidence from Basket Trials: A Bayesian Hierarchical Approach"

**Running title: Bayesian Hierarchical Models for Tumor-Agnostic Therapies**

**Supplementary File**

eTable 1. Estimated intraclass correlation metrics of random-effects models
eTable 2. Sensitivity analysis for modeling objective response rate
eTable 3. Sensitivity analysis for modeling median progression-free survival
eTable 4. Sensitivity analysis for modeling median overall survival

eTable 1. Estimated intraclass correlation metrics of random-effects models

| **Outcome** | **Value** |
| --- | --- |
| Objective response rate | 0.02 |
| Median Progression-free Survival | 0.87 |
| Median Overall Survival | 0.70 |

eTable 2. Sensitivity analysis for modeling objective response rate

| Mean (95%CrL) | Trial | Uninformative prior | | | Weakly informative prior | |
|---|---|---|---|---|---|---|
| | | Base case $\tau$~Uniform(0, 3) | $\tau$~Uniform(0, 5) | $\tau$~Truncnorm(0, 100) | $\tau$~Exp(1) | $\tau$~Exp(0.3) |
| Colorectal | 0.33 | 0.35 (0.25-0.45) | 0.35 (0.25-0.45) | 0.35 (0.25-0.45) | 0.35 (0.26-0.45) | 0.35 (0.25-0.45) |
| Endometrial | 0.48 | 0.44 (0.34-0.55) | 0.44 (0.34-0.55) | 0.44 (0.34-0.55) | 0.43 (0.34-0.54) | 0.44 (0.34-0.55) |
| Gastric | 0.31 | 0.34 (0.22-0.45) | 0.34 (0.22-0.45) | 0.34 (0.22-0.45) | 0.35 (0.23-0.45) | 0.34 (0.22-0.45) |
| Cholangiocarcinoma | 0.41 | 0.39 (0.25-0.54) | 0.39 (0.26-0.54) | 0.39 (0.25-0.54) | 0.38 (0.26-0.53) | 0.39 (0.26-0.53) |
| Pancreatic | 0.18 | 0.3 (0.14-0.43) | 0.3 (0.14-0.43) | 0.3 (0.14-0.43) | 0.31 (0.15-0.44) | 0.3 (0.14-0.43) |
| Small intestine | 0.48 | 0.42 (0.29-0.58) | 0.42 (0.29-0.58) | 0.42 (0.29-0.58) | 0.41 (0.29-0.57) | 0.41 (0.29-0.58) |
| Ovarian | 0.53 | 0.36 (0.22-0.49) | 0.36 (0.22-0.49) | 0.36 (0.22-0.49) | 0.36 (0.23-0.48) | 0.36 (0.22-0.49) |
| Predicted (RE) | | 0.37 (0.15-0.64) | 0.37 (0.15-0.64) | 0.37 (0.15-0.63) | 0.37 (0.18-0.59) | 0.37 (0.16-0.61) |
| Pooled (RE) | | 0.37 (0.26-0.47) | 0.37 (0.26-0.47) | 0.37 (0.26-0.47) | 0.37 (0.27-0.46) | 0.37 (0.27-0.47) |
| Between-tumor SD | | 0.4 (0.02-1.11) | 0.41 (0.03-1.13) | 0.4 (0.03-1.13) | 0.33 (0.02-0.9) | 0.38 (0.03-1.05) |

Abbreviations: RE, random-effects model; SD, standard deviation; Exp, exponential distribution; Truncnorm, truncated normal distribution.

eTable 3. Sensitivity analysis for modeling median progression-free survival

| Mean (95%CrL) | Trial | Likelihood: Lognormal Distribution | | | |
|---|---|---|---|---|---|
| | | Base case $\tau \sim$Uniform(0, 5) | $\tau \sim$Exp(1) | $\tau \sim$Truncnorm(0, 4) | $\tau \sim$Uniform(0, 3) |
| Colorectal | 4.1 | 4.1 (2.87-5.83) | 4.1 (2.88-5.8) | 4.1 (2.9-5.83) | 4.09 (2.87-5.82) |
| Endometrial | 13.1 | 12.7 (9.25-17.41) | 12.66 (9.22-17.33) | 12.69 (9.26-17.36) | 12.73 (9.31-17.5) |
| Gastric | 3.2 | 3.23 (2.11-4.96) | 3.24 (2.11-4.98) | 3.22 (2.1-4.95) | 3.23 (2.1-4.94) |
| Cholangiocarcinoma | 4.2 | 4.18 (2.34-7.42) | 4.18 (2.36-7.41) | 4.16 (2.32-7.39) | 4.16 (2.34-7.5) |
| Pancreatic | 2.1 | 2.21 (1.22-3.95) | 2.23 (1.25-4.01) | 2.21 (1.23-3.97) | 2.21 (1.23-3.97) |
| Small intestine | 23.4 | 20.47 (11.53-35.94) | 20.07 (11.4-35.11) | 20.28 (11.62-35.75) | 20.52 (11.75-35.86) |
| Ovarian | 2.2 | 2.29 (1.3-4.02) | 2.33 (1.33-4.03) | 2.31 (1.31-4.05) | 2.3 (1.31-4.02) |
| Brain | 1.1 | 1.29 (0.6-2.76) | 1.33 (0.62-2.84) | 1.31 (0.61-2.77) | 1.3 (0.6-2.8) |
| Predicted (RE) | | 3.75 (0.24-50.45) | 3.83 (0.37-36.83) | 3.83 (0.28-44.13) | 3.82 (0.25-51.09) |
| Pooled (RE) | | 3.7 (1.46-8.19) | 3.87 (1.7-7.87) | 3.81 (1.55-7.99) | 3.79 (1.51-8.23) |
| Between-tumor SD | | 3.27 (1.87-10.12) | 2.83 (1.8-6.22) | 3.09 (1.85-7.97) | 3.23 (1.86-9.14) |

Abbreviations: RE, random-effects model; SD, standard deviation; Exp, exponential distribution; Truncnorm, truncated normal distribution.

eTable 4. Sensitivity analysis for modeling median overall survival

| **Mean (95%CrL)** | **Trial** | **Likelihood: Lognormal Distribution** | | | |
|---|---|---|---|---|---|
| | | Base case $\tau$~Uniform(0, 5) | $\tau$~Exp(1) | $\tau$~Truncnorm(0, 4) | $\tau$~Uniform(0, 3) |
| Colorectal | 33* | 32.39 (22.78-46.08) | 32.17 (22.58-45.83) | 32.31 (22.67-46.22) | 32.29 (22.67-45.98) |
| Endometrial | 46.5* | 45.34 (33.08-62.2) | 45.19 (32.96-62.13) | 45.33 (33.04-62.48) | 45.36 (32.93-62.35) |
| Gastric | 11.0 | 11.15 (7.2-17.2) | 11.21 (7.29-17.17) | 11.16 (7.22-17.25) | 11.17 (7.27-17.18) |
| Cholangiocarcinoma | 19.4 | 19.15 (10.7-34.37) | 19.1 (10.67-34.02) | 19.07 (10.6-34.34) | 19.1 (10.56-34.65) |
| Pancreatic | 3.7 | 4.1 (2.22-7.49) | 4.21 (2.32-7.65) | 4.13 (2.27-7.52) | 4.09 (2.23-7.49) |
| Small intestine | 36+ | 75.48 (25.57-782.8) | 63.8 (24.69-372.1) | 70.59 (25.36-536) | 73.37 (25.32-620.1) |
| Ovarian | 33.6 | 32.01 (18.2-56.07) | 31.63 (17.88-54.94) | 31.93 (18.28-55.76) | 31.84 (18.08-55.97) |
| Brain | 5.6 | 6.26 (2.91-13.35) | 6.47 (3.04-13.54) | 6.34 (2.95-13.39) | 6.26 (2.92-13.24) |
| Predicted (RE) | | 13.76 (0.42-276.5) | 15.23 (1.13-172.4) | 14.32 (0.72-222) | 14.22 (0.59-254.5) |
| Pooled (RE) | | 13.9 (4.02-34.49) | 15.11 (5.8-32.71) | 14.47 (4.95-33.77) | 14.18 (4.68-33.82) |
| Between-tumor SD | | 3.92 (1.92-19.78) | 3.1 (1.82-8.31) | 3.5 (1.9-11.47) | 3.69 (1.91-12.75) |

Abbreviations: RE, random-effects model; SD, standard deviation; Exp, exponential distribution; Truncnorm, truncated normal distribution.
+, censored at the end of the study period; * Estimated from the fitted exponentiation distribution to the digitized KM curve data.
